# Rapid sepsis diagnosis with protease activity measurement

**DOI:** 10.1101/2025.07.14.25331514

**Authors:** Emily R. Caton, Yezhi Pan, Kiana M. Kiser, Caroline R. Haddaway, Wayne A. Bryden, Michael McLoughlin, Marek A. Mirski, Robert H. Christenson, Cindy C. Sevilla, Sheng Feng, Shuo Chen, Dapeng Chen

**Affiliations:** Zeteo Tech, Inc. Sykesville, MD 21784, USA; Department of Psychiatry, Maryland Psychiatric Research Center, School of Medicine, University of Maryland, Baltimore, MD 21201, USA; Department of Anesthesiology and Critical Care Medicine, Johns Hopkins University School of Medicine, Baltimore, MD 21205, USA; Department of Pathology, University of Maryland School of Medicine, Baltimore, MD 21201, USA; University of Maryland Medical System, Baltimore, MD 21201, USA; Division of Biostatistics and Bioinformatics, Department of Epidemiology and Public Health, School of Medicine, University of Maryland, Baltimore, MD 21205, USA

## Abstract

Human proteases play major roles in various pathological conditions, including dysregulated immune responses in sepsis, making them strong candidates for developing diagnostic markers. Despite this potential, the progress of developing protease-based diagnostic tools has remained slow due to significant technical barriers associated with measuring protease activity, mainly stemming from the vast diversity and the lack of substrate specificity, which complicate the interpretation of protease activity profiles. In this work, we advanced the current state of assay development by designing substrate molecule sensors and implementing an analytical approach based on mass spectrometry. Specifically, we chemically modified protease substrates for human neutrophil elastase (HNE) and matrix metalloproteinases (MMPs) to enhance specificity in mass spectrometry. This approach yields distinct cleavage products with non-overlapping mass-to-charge signatures, allowing precise differentiation of each protease’s activity. We then integrated the modified substrates into a mass spectrometry-based multiplexed assay platform, enabling quantification of multiple protease activities in a single run. We applied the assay to plasma samples and demonstrated that the assay detects distinct protease activity profiles. Our study demonstrated that the assay achieved a diagnostic sensitivity of 88% and specificity of 87% for sepsis detection. The combination of low cost, rapidness, and robust diagnostic performance makes this platform well-suited to a wide range of clinical settings.

**One Sentence Summary:** Novel modifications to protease substrates enable a multiplexed activity assay for accurate sepsis diagnosis in a 3-hour timeframe.

## INTRODUCTION

Sepsis is a life-threatening condition caused by a dysregulated host response to infection, leading to organ dysfunction and, if left untreated, death. Despite advances in critical care, sepsis remains a leading cause of mortality and morbidity worldwide, accounting for an estimated 11 million deaths annually, nearly 20% of global mortality (1). Early diagnosis and swift intervention are critical to improving patient outcomes. Current sepsis diagnostics, ranging from blood cultures and multiplex PCR panels to host-response biomarkers like procalcitonin that aim to detect pathogens or inflammatory signals in the bloodstream. However, these methods often suffer from slow turnaround times, limited sensitivity at low pathogen loads, and poor specificity in non-infectious inflammatory conditions (1). Consequently, there is a critical need for a rapid, sensitive and specific assay for sepsis detection.

Conventional diagnostics for sepsis typically rely on clinical scoring systems (e.g., SOFA, qSOFA), microbial cultures, and non-specific biomarkers such as C-reactive protein (CRP) and procalcitonin (PCT) (2). While blood cultures are the gold standard for pathogen identification, they are often slow (requiring 24 to 72 hours), suffer from low sensitivity, and frequently yield negative results, particularly when patients have received empirical antibiotics. Moreover, non-pathogen-specific biomarkers lack the sensitivity and specificity to distinguish infectious from non-infectious inflammation or stratify patients based on disease severity and progression (2).

Human proteases play a central role in the host immune response during sepsis. These enzymes, particularly neutrophil serine proteases (e.g., human neutrophil elastase [HNE]) and matrix metalloproteinases (e.g., MMP8 and MMP9), are released during neutrophil activation and participate in tissue remodeling, microbial clearance, and inflammatory signaling (3). However, in sepsis, their activities can become dysregulated, contributing to excessive tissue damage, vascular leakage, and multiorgan failure (4). Importantly, their activity levels, not just their abundance, may offer real-time insight into the pathological processes of sepsis, including the onset and severity of the condition. Leveraging protease activity as a diagnostic marker offers a novel and potentially transformative approach to sepsis detection. Unlike conventional protein biomarkers that reflect gene expression or immune activation in a delayed fashion, protease activity provides an immediate readout of enzymatic function and inflammatory dynamics (5). Since protease dysregulation occurs early in sepsis pathophysiology, measuring specific protease activities enables a faster and mechanistically informative diagnosis. Moreover, our functional assay targeting protease activity inherently differs from traditional antigen or nucleic acid-based diagnostics as it captures dynamic biological processes which may better correlate with clinical trajectory and therapeutic response (6).

Leveraging the pivotal role of human proteases in sepsis, we developed an innovative approach that modifies protease substrates to enhance specificity and simultaneously quantifies multiple protease activities in a single multiplexed assay (7). We applied our multiplexed assay targeting HNE and MMPs to sepsis-patient plasma. Our assay reliably distinguished distinct protease activity signatures and achieved excellent diagnostic performance with a fast turnaround time. Given its rapid turnaround time, low operational cost, and high specificity, this platform has the potential to substantially transform current sepsis-diagnostic practices.

## RESULTS

### Concept of the substrate modification for the multiplexed protease activity assay

Our approach of designing a new type of protease substrates derived from conventionally reported peptide sequences cleaved by human proteases. To enhance the analytical capacity of these substrates, we conjugate each peptide to a polymer molecule, primarily polyethylene glycol (PEG) (Fig 1A). Upon exposure to human proteases, the polymer-linked substrate is cleaved, but the reaction is broadly specific due to the enzyme’s nonspecific activity and their diversity and abundance in human specimens (Fig 1A). Consequently, cleavage occurs at multiple potential sites, generating a heterogeneous mixture of peptide fragments (Fig. 1B). We use matrix-assisted laser desorption/ionization time-of-flight mass spectrometry (MALDI-TOF MS) to analyze the resulting cleavage products, accurately revealing the cleavage sites and the protease activity profile (Fig. 1B). The polymer linker also allows us to fine-tune the molecular weight of each substrate so that each produces a distinct, non-overlapping mass peak pattern in the mass spectrum (Fig 1C). This unique strategy allows us to include multiple substrates in a single assay, allowing multiplexed detection of protease activities within one MALDI-TOF MS run (Fig 1C).

**Fig. 1.**
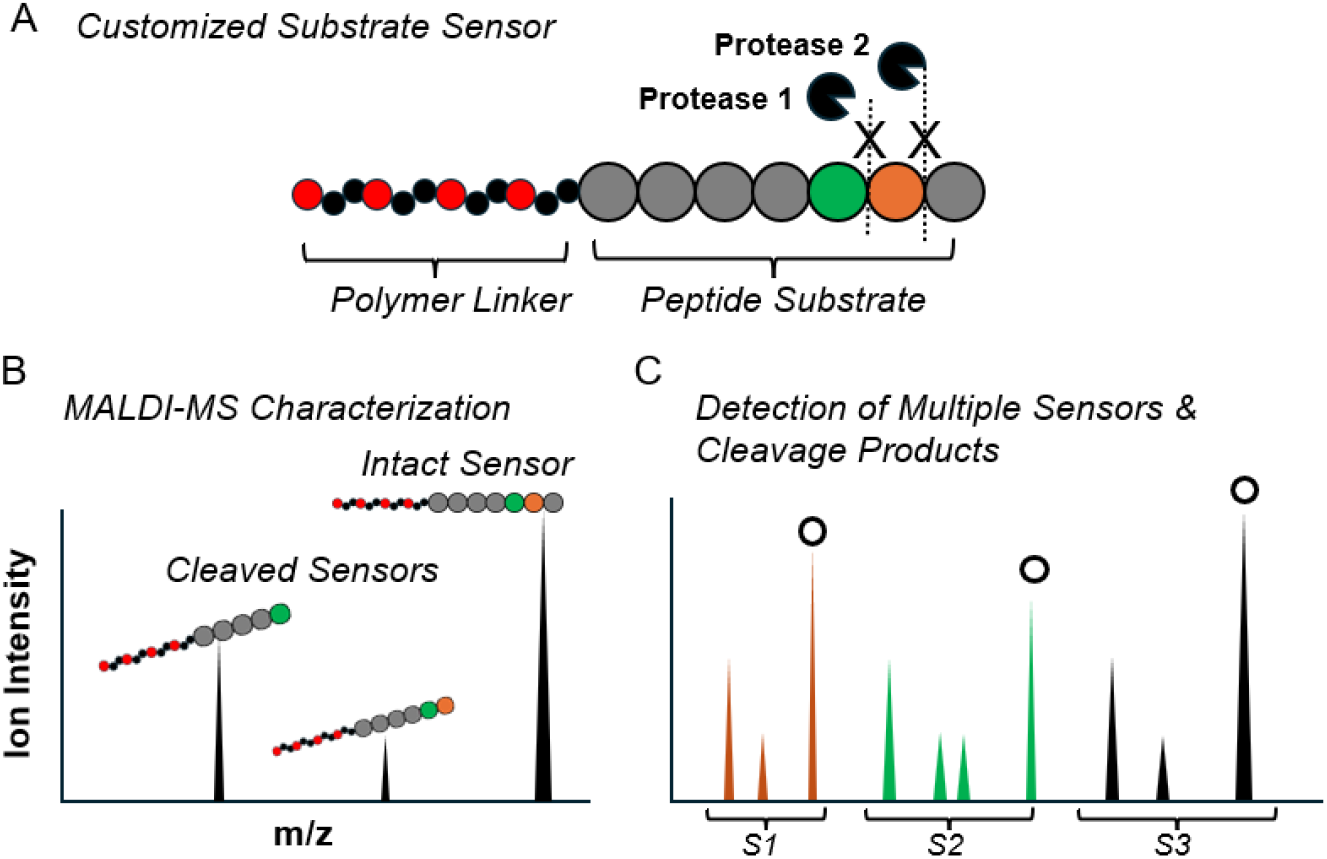
Overview of the substrate sensor design and the activity measurement of multiple proteases based on mass spectrometry. **(A)** The development of customized substrates sensors. **(B)** The detection of protease activity using the customized sensor with mass spectrometry. **(C)** The detection of multiple protease activity using customized substrate sensors and mass spectrometry.

### Multiplexed protease activity assay development

We have designed and implemented three customized substrate sensors for our multiplexed protease activity assay by modifying peptide substrates for HNE, ES001, and ES010 (S1, S2, and S3) (Fig. 2A, B and C). To differentiate these substrate sensors clearly in MALDI-TOF MS, we conjugated PEG molecules of different lengths for distinct separation based on mass differences (Fig. 2C). Additionally, to minimize the interference from the common presence of detergents in protease assays, we increased the molecular weight of each substrate sensor to exceed 2000 m/z to reduce background interference during mass spectrometric detection (Fig. 2A). Consequently, when analyzed by MALDI-TOF MS, the substrate sensors showed clear signals without background noise (Fig. 2A). It is noteworthy that individual substrate sensors typically do not show as a single mass peak in MALDI-TOF mass spectrometry; rather, they often appear as clusters of sodium adducts due to the inherent nature of their peptide sequences (Fig. 2A). However, these sodium adduct peaks are highly consistent and correlate strongly with the primary peptide signals. Subsequent incubation of the three substrate sensors with plasma specimens revealed distinct cleavage patterns. Specifically, S1 produced one identifiable cleavage product (S1-1); S2 produced four separate cleavage fragments (S2-1 to S2-4); and S3 produced two fragments (S3-1 and S3-2) (Fig. 2B).

**Fig. 2.**
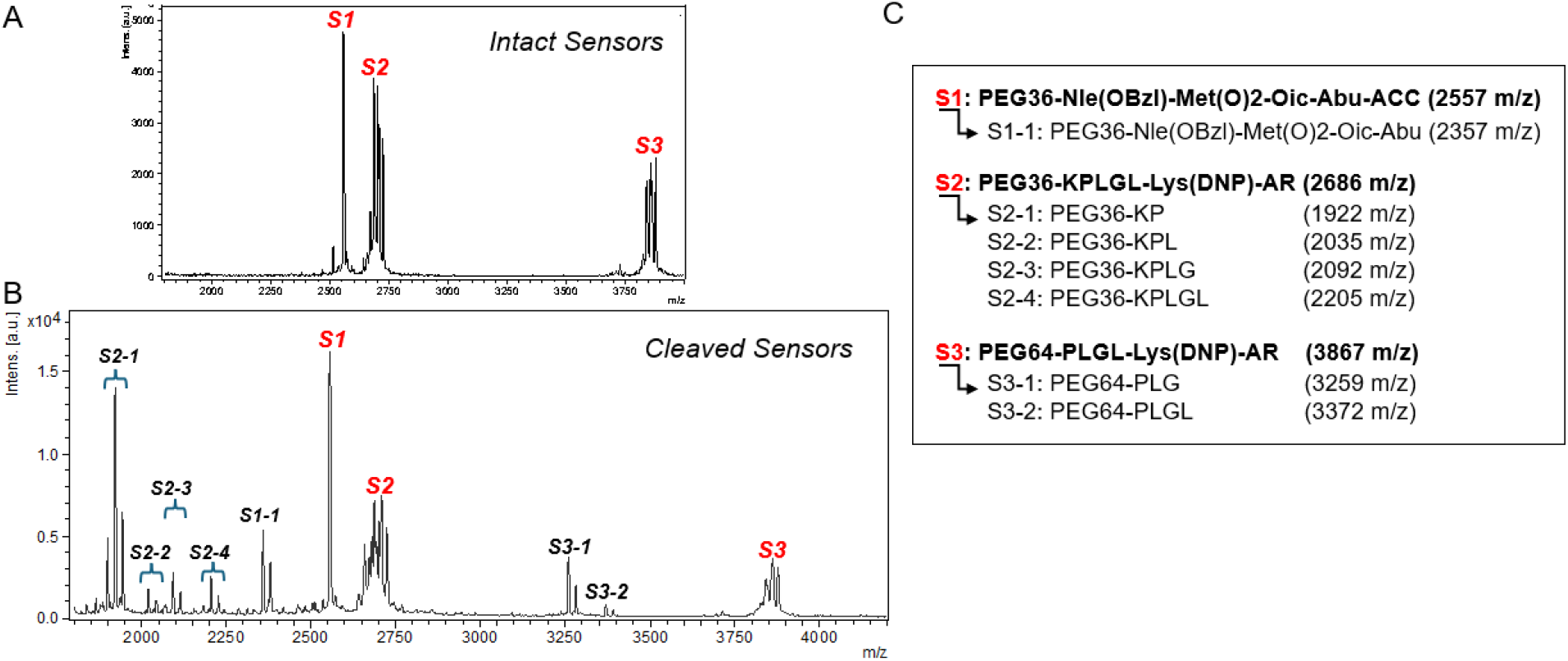
Overview of the customized substrate sensors for HNE and MMPs in the multiplexed assay. **(A)** The three protease substrate sensors used in the multiplexed protease activity assay. **(B)** The demonstration of intact substrate sensors and their cleavage products upon the incubation with plasma. **(C)** Three substate sensors and the cleavage product information.

The cleavage mechanism for S1 has been extensively studied in our prior studies and is well characterized, specifically involving the removal of the ACC group by HNE. In this study, accurate mass measurements demonstrated sequential cleavage of sensor S2, beginning from leucine to proline at the C-terminal, with each amino acid cleaved in an orderly manner (Fig. 2C). A similar sequential cleavage pattern was also observed in S3 (Fig. 2C). Furthermore, since each of these resulting fragments exhibited varying peak intensities in the mass spectra, protease activities can be quantified by analyzing the intensity of patterns associated with relevant cleavage products (Fig. 2B).

### Substrate sensor profiles in sepsis and non-sepsis patients

To evaluate the clinical utility of our multiplexed protease activity assay, we obtained plasma from sepsis patients (n = 47) and non-sepsis controls (n = 40) from the University of Maryland Medical Center (UMMC) Core Laboratory and analyzed their protease activity profiles (Table 1). The cohort exhibited a broad spectrum of acute presentations, including fever, altered mental status, unresponsiveness, abdominal pain, breast mass, back pain, anemia, chemotherapy-related complications, transfusion requirements, and trauma. The clinical setting are also diversified, reflecting multiple healthcare settings within the Maryland healthcare system, including emergency departments, general inpatient medical and surgical units, and specialized intensive care units (ICUs).

**Table 1.** Patient demographic data and substrate sensors profiles in the multiplexed protease activity assay.

| Variable | Sepsis (n= 47 ) | Non-Sepsis (n= 40 ) | p-value |
| --- | --- | --- | --- |
| Age | 58.27 (15.13) | 58.07 (22.69) | 0.965 |
| Gender | 23 (50%) | 18 (47%) | 0.964 |
| Lactate | 2.81 (1.9) | 2.55 (2.99) | 0.679 |
| S1-1 | 0.27 (0.34) | 0.54 (1.8) | 0.426 |
| S2-1 | 1 (1.61) | 5.35 (6.31) | <0.001 |
| S2-2 | 0.25 (0.43) | 3.63 (4.65) | <0.001 |
| S2-3 | 0.97 (1.35) | 1.88 (1.04) | 0.002 |
| S2-4 | 0.69 (1.18) | 3.74 (3.6) | <0.001 |
| S3-1 | 0.8 (0.84) | 3.83 (3.8) | <0.001 |
| S3-2 | 0.3 (0.51) | 1.61 (1.59) | <0.001 |

There were no statistically significant differences in age or gender distribution between sepsis and non-sepsis groups (Table 1). Additionally, the whole blood lactate levels from blood gas analyzer, did not differ significantly between the two groups (Table 1), underscoring the whole-blood lactate’s limited specificity as a standalone sepsis biomarker. Our results showed that S1-1 did not have significant differences between patient groups (Table 1, Fig. 3A). However, significant differences were observed in the profiles of S2 and S3 (Table 1, Fig. 3B-F).

**Fig. 3.**
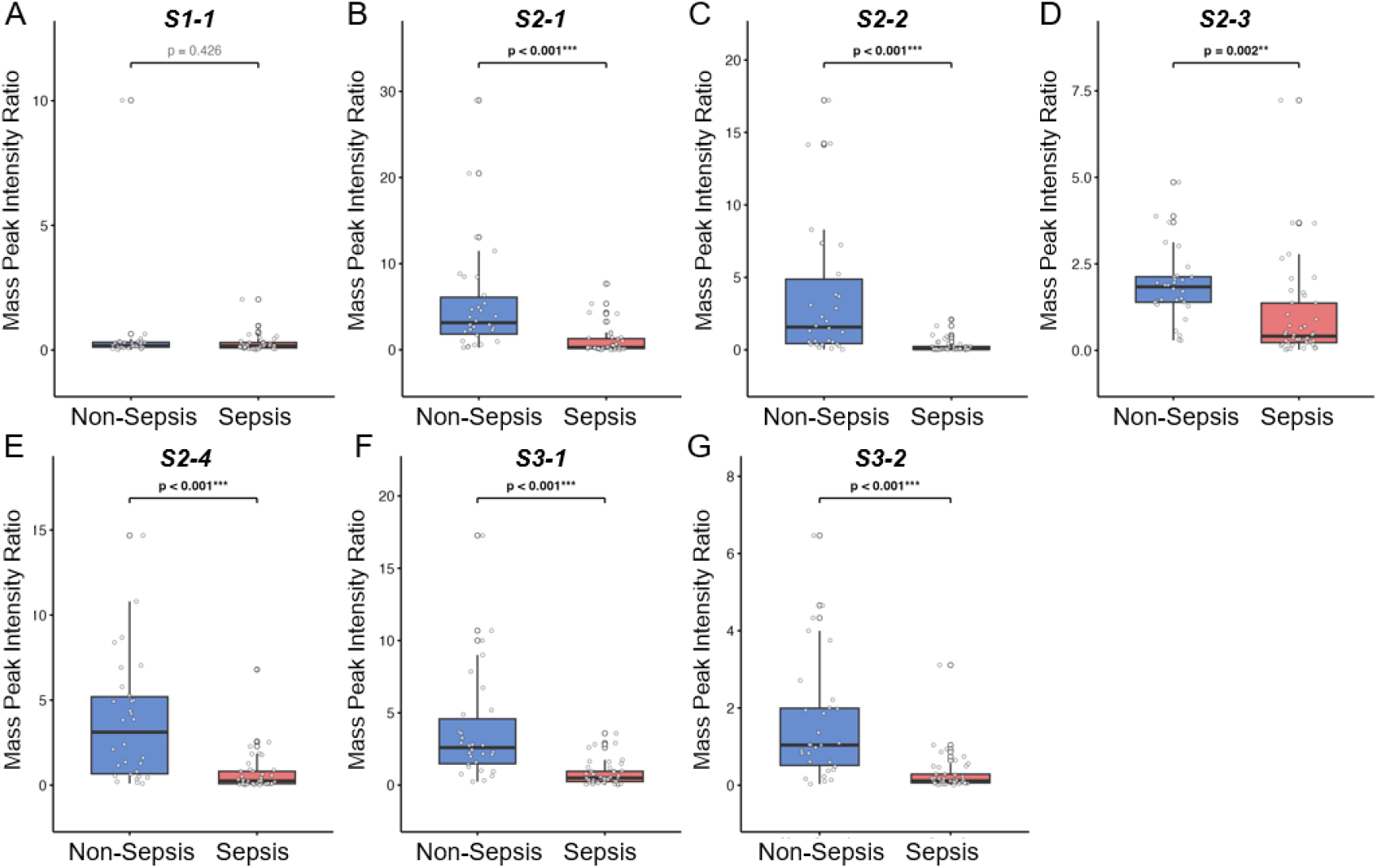
Substrate sensor profiles for the multiplexed protease activity assay in sepsis and non-sepsis patients. **(A)** S1-1 profile in sepsis and non-sepsis patients. **(B)** S2-1 profile in sepsis and non-sepsis patients. **(C)** S2-2 profile in sepsis and non-sepsis patients. **(D)** S2-3 profile in sepsis and non-sepsis patients. **(E)** S2-4 profile in sepsis and non-sepsis patients. **(F)** S3-1 profile in sepsis and non-sepsis patients. **(G)** S3-2 profile in sepsis and non-sepsis patients. The data are presented using box-and-whisker plots, where each box indicates the interquartile range with the median line, whiskers extend to 1.5x IQR, and individual points show raw data. Independent t-tests were used for group comparisons, with two-sided p-values annotated above each plot.

Specifically, the ratio of S2-1 to S3-2 was significantly lower in non-sepsis plasma than in sepsis plasma (Table 1, Fig. 3B-F). This finding suggests that protease activity is elevated in patients with sepsis, demonstrating the assay’s robust ability to differentiate sepsis from non-sepsis.

Continuous variables are presented as mean values accompanied by standard deviations (SD), while categorical variables are reported as counts with corresponding percentages. Statistical comparisons between groups were conducted using independent t-tests for continuous variables, and chi-square tests were used for categorical variables. All p-values were two-sided, with statistical significance set at p < 0.05.

### The multiplexed protease activity assay shows great diagnosis accuracy for sepsis

The significant differences observed in the S2 and S3 substrate sensor profiles between sepsis and non-sepsis patient groups indicated the potential use of these markers for sepsis diagnosis. We employed the XGBoost to develop predictive models for sepsis classification using various data sets, including whole blood lactate measurements alone, individual substrate sensor profiles separately, and the combined data from all three substrate sensors obtained over a three-hour incubation (Table 2). Consistent with our expectations, combining all three substrate sensors achieved the highest diagnostic performance among all tested strategies (Table 2, Fig. 4A-E). Specifically, the combined sensor model yields an accuracy of 87.5%, sensitivity of 88%, specificity of 86.7%, positive predictive value (PPV) of 92.0%, negative predictive value (NPV) of 88.7%, recall of 88.0%, and an area under the receiver operating characteristic curve (AUC) of 0.823 (Table 2, Fig. A-E). These results suggest combing multiple substrate sensor profiles delivers superior diagnostic performance compared to individual markers or conventional lactate testing alone.

**Table 2.**
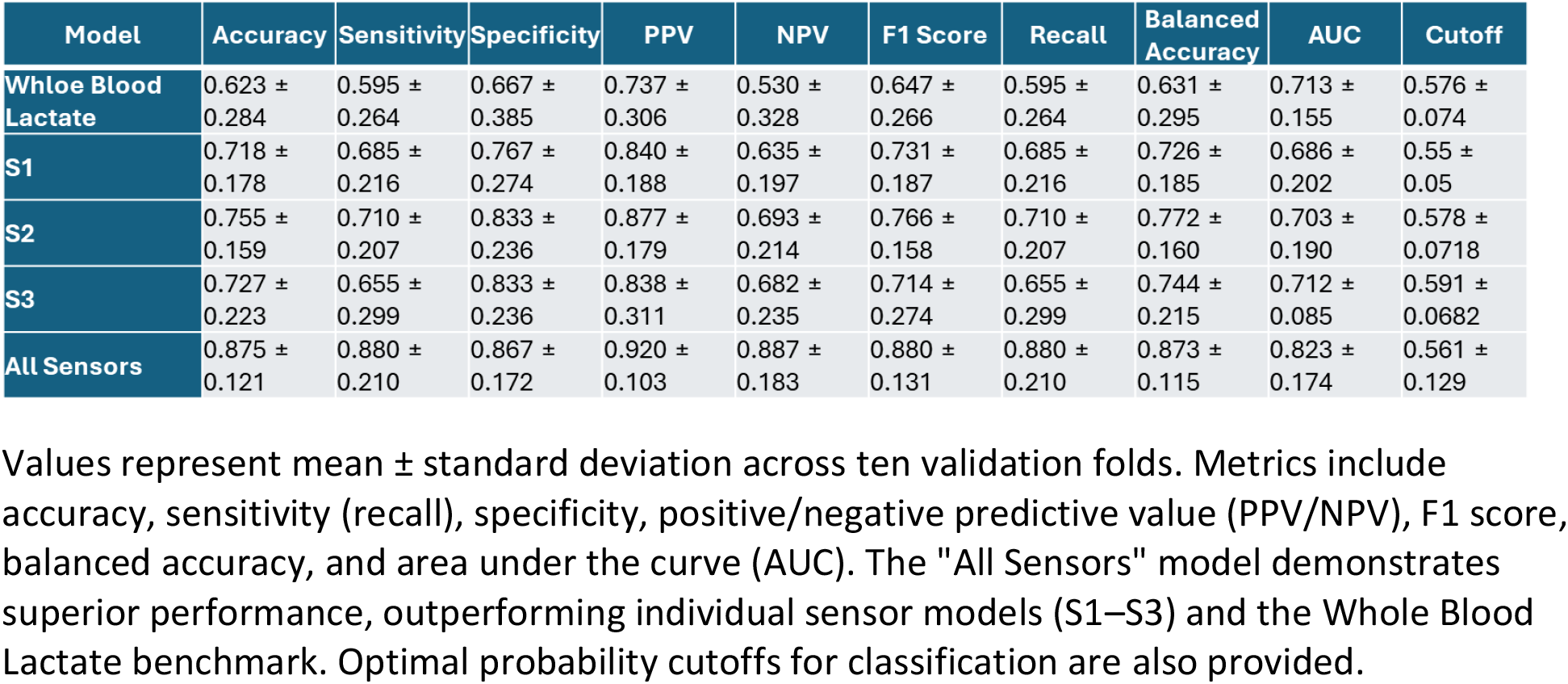
Performance matrices.

| Model | Accuracy | Sensitivity | Specificity | PPV | NPV | F1 Score | Recall | Balanced Accuracy | AUC | Cutoff |
| --- | --- | --- | --- | --- | --- | --- | --- | --- | --- | --- |
| Whole Blood Lactate | 0.623 ± 0.284 | 0.595 ± 0.264 | 0.667 ± 0.385 | 0.737 ± 0.306 | 0.530 ± 0.328 | 0.647 ± 0.266 | 0.595 ± 0.264 | 0.631 ± 0.295 | 0.713 ± 0.155 | 0.576 ± 0.074 |
| S1 | 0.718 ± 0.178 | 0.685 ± 0.216 | 0.767 ± 0.274 | 0.840 ± 0.188 | 0.635 ± 0.197 | 0.731 ± 0.187 | 0.685 ± 0.216 | 0.726 ± 0.185 | 0.686 ± 0.202 | 0.55 ± 0.05 |
| S2 | 0.755 ± 0.159 | 0.710 ± 0.207 | 0.833 ± 0.236 | 0.877 ± 0.179 | 0.693 ± 0.214 | 0.766 ± 0.158 | 0.710 ± 0.207 | 0.772 ± 0.160 | 0.703 ± 0.190 | 0.578 ± 0.0718 |
| S3 | 0.727 ± 0.223 | 0.655 ± 0.299 | 0.833 ± 0.236 | 0.838 ± 0.311 | 0.682 ± 0.235 | 0.714 ± 0.274 | 0.655 ± 0.299 | 0.744 ± 0.215 | 0.712 ± 0.085 | 0.591 ± 0.0682 |
| All Sensors | 0.875 ± 0.121 | 0.880 ± 0.210 | 0.867 ± 0.172 | 0.920 ± 0.103 | 0.887 ± 0.183 | 0.880 ± 0.131 | 0.880 ± 0.210 | 0.873 ± 0.115 | 0.823 ± 0.174 | 0.561 ± 0.129 |

**Fig. 4.**
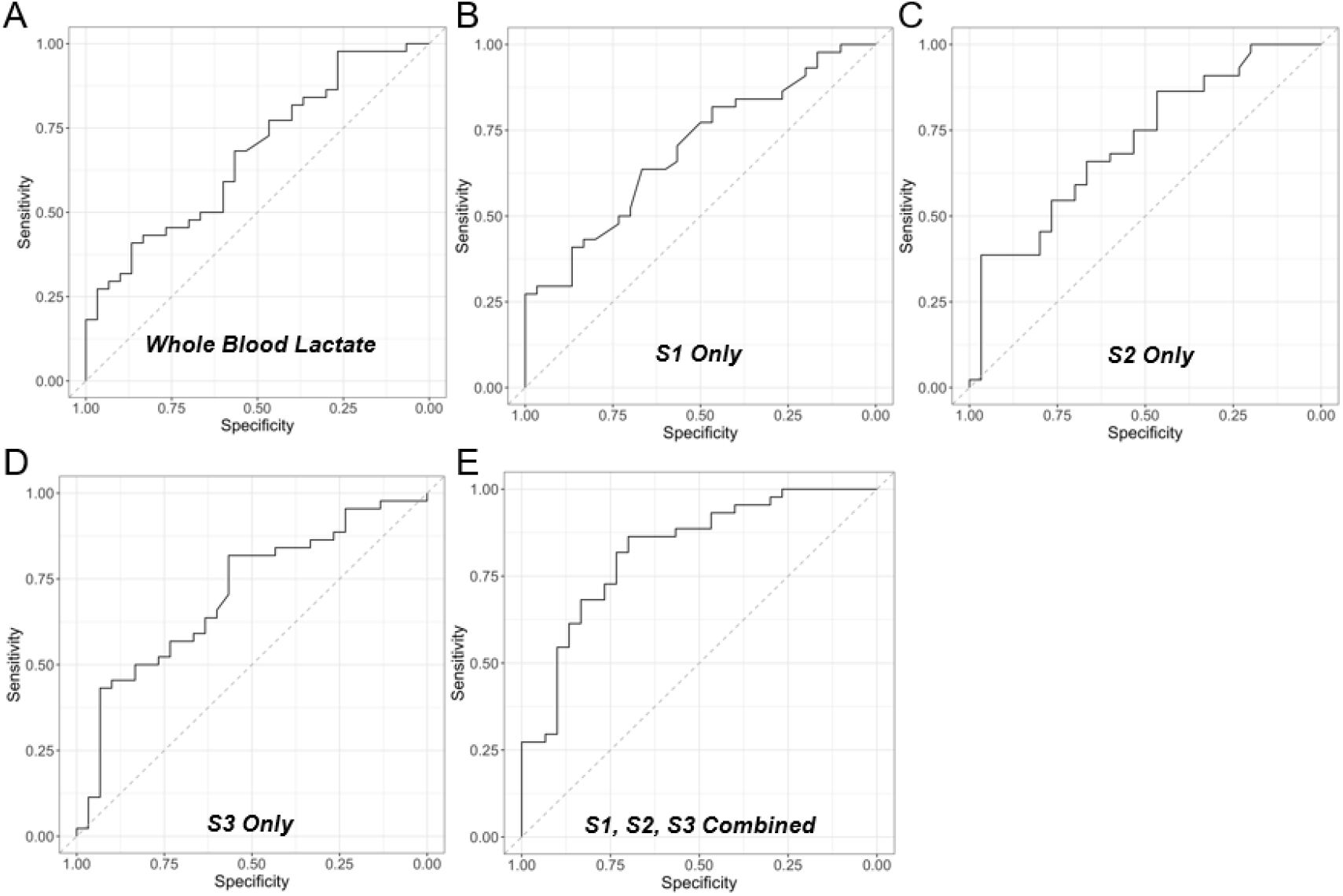
ROC of whole blood lactate and substrate sensor profiles in the multiplexed protease activity assay for distinguishing between sepsis and non-sepsis. **(A)** ROC curve of whole blood lactate for differentiating sepsis versus non-sepsis patients. **(B)** ROC curve of S1 profiles acquired from the multiplexed protease activity assay lactate for differentiating sepsis versus non-sepsis patients. **(C)** ROC curve of S2 profiles acquired from the multiplexed protease activity assay for differentiating sepsis versus non-sepsis patients. **(D)** ROC curve of S3 profiles acquired from the multiplexed protease activity assay for differentiating sepsis versus non-sepsis patients. **(E)** ROC curve of combining all three substrate sensor profiles acquired from the multiplexed protease activity assay for differentiating sepsis versus non-sepsis patients.

Values represent mean ± standard deviation across ten validation folds. Metrics include accuracy, sensitivity (recall), specificity, positive/negative predictive value (PPV/NPV), F1 score, balanced accuracy, and area under the curve (AUC). The “All Sensors” model demonstrates superior performance, outperforming individual sensor models (S1–S3) and the Whole Blood Lactate benchmark. Optimal probability cutoffs for classification are also provided.

## DISCUSSION

Our multiplexed protease activity assay, using customized substrate sensors coupled with MALDI-TOF MS, represents a substantial advancement in the application of protease activity profiling for clinical diagnostic. Conventional fluorescence-based protease assays are limited, as they integrate signals from multiple cleavage events generated by different proteases into a single readout, preventing discrimination of individual protease activities. In contrast, our MALDI-TOF MS coupled method employs customized PEG-linker substrates bearing unique mass tags, which generate distinct protease-specific cleavage products that can be individually resolved and quantified in a single reaction, thereby significantly improving assay specificity and achieving genuine multiplexing. Such multiplexed profiling is particularly valuable for diagnostic applications where differentiation among proteases with overlapping substrate specificities is required in clinical samples. The integration of PEG-linker substrates with mass-based detection represents a novel approach in the field of protease diagnostics, this specific combination has not been previously demonstrated either in academic studies or clinical assay development.

Sepsis remains a critical global health burden, demanding rapid and accurate diagnosis to guide early interventions and improve patient outcomes. Despite growing awareness and clinical emphasis on early sepsis recognition, current diagnostic approaches are still facing significant challenges, including prolonged turnaround times, suboptimal specificity, high specimen volume requirements, and dependency on resource-intensive laboratory infrastructure, leading to underdiagnosis, misdiagnosis, and delays in initiating life-saving therapies such as appropriate antimicrobial treatment or supportive care.

Procalcitonin previously has been considered as a promising tirage marker for sepsis. However the current procalcitonin assay based on antibody-antigen recognition often suffers interference from hemolysis (Influence of hemolysis on serum procalcitonin measured by electrochemiluminescence in an emergency room), lipemia (Multicenter evaluation of a new immunoassay for procalcitonin measurement on the Kryptor System), and biotin supplementation (Interference From High-Dose Biotin Intake in Immunoassays for Potentially Time-Critical Analytes by Roche: Evaluation of a Countermeasure for Worst-Case Scenarios). Reported sensitivity and specificity for procalcitonin typically range from 65%-80%, depending on the cutoff and patient population. Recent FDA-cleared sepsis diagnostic tools including IntelliSep (AUC 0.74), Septicyte® Rapid (AUC 0.71 – 0.80), Sepsis ImmunoScore™ (AUC 0.80-0.85) have improved performance. Although Sepsis ImmunoScore™ showed great diagnostic performance, this AI-powered diagnostic tool depends on 22 predetermined patient parameters including demographic data, vital sign measurements complete metabolic panel, complete blood count panel, lactate level, procalcitonin, and CRP levels, and may be vulnerable to analytical interference and significant infrastructure dependencies. In contrast, our multiplexed protease assay achieved a great diagnostic performance (AUC of ROC 0.823) with a simplified workflow and minimal sample volume.

The platform described in this study addresses these challenges by offering a rapid, sensitive, and low-resource diagnostic solution for sepsis based on multiplexed human protease activity profiling. When applied to patient plasma, our assay demonstrated strong diagnostic performance for sepsis with an accuracy of 87.5%, sensitivity of 88.0%, specificity of 86.7%, and an AUC of 0.823, alongside a PPV of 92.0% and NPV of 88.7%, underscoring its reliability for both ruling in and ruling out sepsis. The high sensitivity and NPV show its value as an effective early rule-out tool, while the high PPV supports decision-making in high-risk patients. Notably, the platform provides these results within a 3-hour turnaround time, using less than 50 microliters of plasma, which represents a significant improvement over conventional diagnostic assays in terms of speed, invasiveness, and operational simplicity. The ability to deliver rapid and accurate results with minimal sample volume and infrastructure requirements positions this platform as a promising solution for point-of-care or decentralized diagnostic workflows.

By facilitating earlier detection of sepsis, this technology can improve patient triage, support more targeted antimicrobial therapy, enable faster escalation of care, and reduced mortality, particularly in settings where delays in diagnosis have been shown to significantly worsen patient outcomes. Given the platform’s high performance and practical advantages, it has the potential to fill a critical gap in current sepsis diagnostic practice and to serve as a foundation for broader application to other inflammatory and infectious disease.

## MATERIALS AND METHODS

### Study design

We initially developed a multiplexed protease activity assay and conducted its analytical evaluation using a panel of recombinant human proteases to confirm the assay’s capability to detect and distinguish individual protease activities. Following this validation, we applied the assay to plasma specimens from patients with or without sepsis to assess the clinical diagnostic performance of the assay in differentiating septic versus non-septic patients based on distinct protease activity profiles.

### Clinical subjects

Plasma specimens were obtained from the core laboratory of the University of Maryland Medical Center (UMMC) during January to February 2025, for whom arterial lactic acid were ordered. All specimens were de-identified and frozen at −20°C after clinical testing was completed, until time of analysis using multiplexed protease activity assay. The study was approved by the Institutional Review Borad at University of Maryland. The classification of specimens into sepsis versus non-sepsis was determined retrospectively by reviewing patients’ medical records. The diagnosis of sepsis, where applicable, was based on the institutional clinical judgment and documentation at the time of patient specimens tested.

### Substrate sensor development

All three substrate sensors were designed by Zeteo Tech and synthesized by CPC Scientific Inc. The synthesized sensors were prepared in high-performance liquid chromatography (HPLC)-grade water with a concentration of 200 µM upon arrival.

### Incubation of substrate sensors with recombinant human proteases

Three substrate sensors with the concentration of 10 µM prepared in water were prepared in a buffer containing sodium salts and detergent. Commercial recombinant human proteases from R&D Systems (Minneapolis, MN) and, including human neutrophil elastase (rhNE) (Catalog # 9167-SE), recombinant human MMP-9 protein (catalog #: 911-MP), and recombinant human MMP-8 protein (catalog #: 908-MP), were mixed with the substrate sensor buffer at 37oC for 10 minutes.

#### Incubation of substrate sensors with plasma

The buffer containing the three substrate sensors was mixed with 6 µL of plasma and incubated at 37oC for either 3 or 24 hours.

### MALDI-TOF MS

For MALDI-TOF MS analysis, 1 µL of each prepared sample was carefully dispensed onto a spot on a Bruker MALDI target plate and allowed to air-dry at room temperature. Following the initial drying, 1 µL of α-cyano-4-hydroxycinnamic acid matrix solution (prepared at a concentration of 9 mg/mL in 70% acetonitrile) was added directly onto the dried sample spot and subsequently dried under the same conditions. The resulting sample-matrix co-crystals were subjected to MALDI-TOF mass spectrometry using a Bruker Daltonics microflex LRF instrument (Billerica, MA), operating in positive ion linear mode. Spectral acquisition was conducted by averaging 500 laser shots to ensure adequate signal intensity and reproducibility, targeting a mass-to-charge (m/z) range of 1,500 to 3,000. Ion intensity values corresponding to the target mass peaks were extracted from the raw data files produced by the Bruker instrument’s proprietary operating software for subsequent analysis.

### Statistical analysis

Descriptive and comparative analyses were conducted to characterize the dataset and evaluate differences between sepsis and non-sepsis groups. Continuous variables are reported as mean ± standard deviation (SD) and compared using independent t-tests. Categorical variables are presented as frequencies (%) and compared using chi-square tests. Boxplots visualize distributions of substrate sensor profiles across groups, with whiskers extending to 1.5× interquartile range (IQR) and individual data points overlaid. Statistical comparisons between groups were annotated with two-sided p-values. All analyses were performed in R (version 4.3.0), with statistical significance set at α = 0.05.

We utilized the Extreme Gradient Boosting (XGBoost) algorithm to train a predictive model for sepsis diagnosis. Each sample was represented as a seven-dimensional feature vector, corresponding to quantitative readouts from seven substrate sensors. The model was trained to classify patients as either sepsis or non-sepsis based on the measurements of the seven sensors. To assess the performance of the classifier, we implemented a 10-fold cross-validation scheme. The dataset was randomly partitioned into ten folds. For each iteration, the model was trained on 90% of the data and evaluated on the remaining 10%. Performance metrics from each fold were then averaged to obtain overall cross-validated estimates. The model outputted a continuous probability score (ranging from 0 to 1) for each sample, reflecting the predicted probability of sepsis. To convert probability scores into binary diagnostic classifications, we determined an optimal probability threshold for each fold using Youden’s J statistic, defined as:

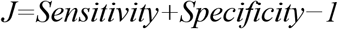

The optimal threshold for each fold was identified by computing the ROC curve and selecting the threshold that maximized Youden’s J statistic. This was implemented using the coords() function in the R package pROC, which internally evaluates sensitivity and specificity across the full range of possible thresholds. The fold-specific optimal cutoff was then applied to binarize the predicted probabilities into sepsis vs. non-sepsis classifications. For each fold, the following metrics were computed based on the respective optimal threshold: accuracy, sensitivity, specificity, and area under the receiver operating characteristic curve (AUC). These metrics were averaged across ten folds to yield the model’s overall cross-validated performance.

## Data Availability

All data are available in the main text or the supplementary materials.

## List of Supplementary Materials

None.

## Acknowledgments

None.

## Author contributions

Conceptualization: DC, MAM

Methodology: DC, SF, SC, RHC

Investigation: ERC, YP, KMK, CRH, CCS

Visualization: DC, YP, SC

Funding acquisition: DC, WAB, MM

Project administration: DC, MM

Supervision: DC, SF, SC

Writing – original draft: DC, SF, SC, YP, WAB, MAM, MM

Writing – review & editing: DC

## Data and materials availability

All data are available in the main text or the supplementary materials.

